# Evidence-based XAI of clinical decision support systems for differential diagnosis: Design, implementation, and evaluation

**DOI:** 10.1101/2024.07.18.24310609

**Authors:** Yasuhiko Miyachi, Osamu Ishii, Keijiro Torigoe

## Abstract

**Introduction:** We propose the Explainable AI (XAI) model for Clinical Decision Support Systems (CDSSs). It supports physician’s Differential Diagnosis (DDx) with Evidence-based Medicine (EBM). It identifies instances of the case data contributing to predicted diseases. Each case data is linked to the sourced medical literature. Therefore, this model can provide medical professionals with evidence of predicted diseases.

**Methods:** The source of the case data (training data) is medical literature. The prediction model (the main model) uses Neural Network (NN) + Learning To Rank (LTR). Physicians’ DDx and machines’ LTR are remarkably similar. The XAI model (the surrogate model) uses k-Nearest Neighbors Surrogate model (k-NN Surrogate model). The k-NN Surrogate model is a symphony of Example-based explanations, Local surrogate model, and k-Nearest Neighbors (k-NN). Requirements of the XAI for CDSS and features of the XAI model are remarkably adaptable. To improve the surrogate model’s performance, it performs “Selecting its data closest to the main model.” We evaluated the prediction and XAI performance of the models.

**Results:** With the effect of “Selecting,” the surrogate model’s prediction and XAI performances are higher than those of the “standalone” surrogate model.

**Conclusions:** The k-NN Surrogate model is a useful XAI model for CDSS. For CDSSs with similar aims and features, the k-NN Surrogate model is helpful and easy to implement. The k-NN Surrogate model is an Evidence-based XAI for CDSSs.

Unlike current commercial Large Language Models (LLMs), Our CDSS shows evidence of predicted diseases to medical professionals.

## Introduction

### Clinical Decision Support System

Clinical Decision Support Systems (CDSSs) aim to improve medical care quality. CDSSs support medical decisions with clinical knowledge, patient information, and other medical information. (1)

The summaries of this article’s CDSS are as follows:

– Objectives

– To support Differential Diagnosis (DDx) by physicians and general practitioners.
– To prevent diagnostic errors for medical professionals.
– To reduce diagnostic uncertainty
– Medical literature, metadata, and case data

Medical literature (textbooks, original articles, case reports, etc.) is a source of metadata and case data.
Metadata (bibliographic and disease-related information) is obtained from medical literature by text-mining.
Case data (symptoms and diseases) is obtained from the metadata by coding. It is used for training and test of the models.
Symptoms include signs and symptoms, laboratory and imaging test results, etc. (defined as “symptoms” from now on).
Diseases include confirmed diseases, differential diseases, and their scores.
– Prediction model

A. medical professional inputs the patient’s symptoms.
B. CDSS outputs predicted diseases by Artificial Intelligence (AI).
C. diseases are a ranking list of diseases. (2)
– XAI model

The CDSS outputs evidence of predicted diseases by Explainable AI (XAI).
Evidence is the metadata of the case data contributing to predicted diseases. (3), (4)

Our CDSS is open to medical professionals on the Internet. (5) (See: Fig 1, S Table 1)

**Fig 1.**
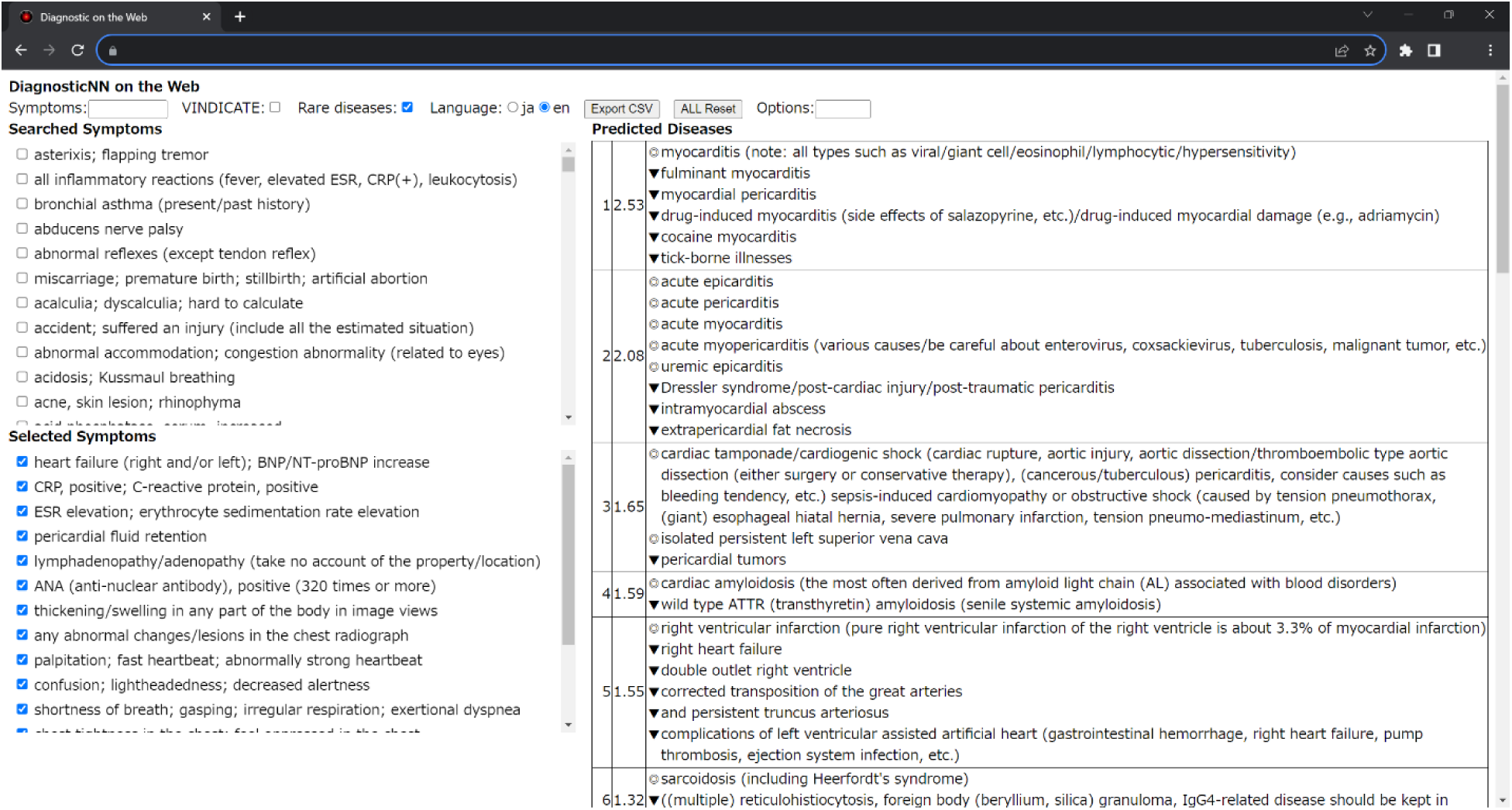
Screen image of our Clinical Decision Support System Case citation: (6)

**Table 1.** Definitions of data.

| Variables, Functions, etc. | Definitions |
| --- | --- |
| Case data |  |
| $X_{train}$ | Training data for features |
| $y_{train}$ | Training data for targets |
| $X_{test}$ | Test data for features |
| $y_{test}$ ($y_{true}$) | Test data for targets (True data for targets) |
| User's input |  |
| $X_{in}$ | User's Inputted data |
| Main model |  |
| $y_{pred} = main\_model.predict(X)$ | The main model's predicted data |
| $ndcg_{nn} = ndcg(y_{true}, y_{pred})$ | The main model's prediction performance |
| Surrogate model |  |
| $k = 1 - 10$ | The number of neighbors |
| $y_{pred\_k[]} = surrogate\_model.predict(X, k)$ | The surrogate model's predicted data |
| $ndcg\_k1[] = ndcg(y_{true}, y_{pred\_k[]})$ | The surrogate model's prediction performance |
| $ndcg\_k2[] = ndcg(y_{pred}, y_{pred\_k[]})$ | The surrogate model's XAI performance |
| $k\_closest = argmax(ndcg\_k2[]) + 1$ | Selecting the surrogate model's predicted data closest to the main model |
| $ndcg\_k1\_closest = ndcg\_k1[k\_closest - 1]$ | The surrogate model's prediction performance with "Selecting" |
| $ndcg\_k2\_closest = ndcg\_k2[k\_closest - 1]$ | The surrogate model's XAI performance with "Selecting" |

[]: Array starting from 0

CDSSs with similar aims and features are available worldwide. (7), (8), (9), (10)

Clinical Decision Support System and Learning To Rank

The prediction model of our CDSS uses Neural Network (NN) + Learning To Rank (LTR).

Physicians’ DDx and machines’ LTR are remarkably similar. (2)

### Explainable AI

Explainable AI (XAI) aims to help humans accept the AI’s behaviors.

The key points are interpretability and explainability. (11)

XAI has a variety of methods. (12), (13), (See: S Table 2)

**Table 2.** Example of metadata and case data.

| Case ID | C##### |  |  |  |
| --- | --- | --- | --- | --- |
| Metadata |  | Case data |  |  |
| Bibliographic |  |  |  |  |
| Authors | Stern S, Cifu A, Altkorn D |  |  |  |
| Title | ***Symptom to Diagnosis: An Evidence-Based Guide*** |  |  |  |
| ISBN-13 | 978-1260121117 |  |  |  |
| Pages | 1 - 8 |  | code | value |
| Disease-related |  |  |  |  |
| **Confirmed diseases** | **Soft tissue: Cellulitis** | → y | d### | 10.0 + $\alpha$ |
| Differential diseases | Skin: Stasis dermatitis |  | d### | 10.0 |
|  | Calf veins: Distal DVT |  | d### | 10.0 |
| Symptoms | Painful left leg edema with erythema | → X | s### | 1 |
|  | Hypertension |  | s### | 1 |
|  | Osteoarthritis of the knees |  | s### | 1 |
|  | Status post cholecystectomy |  | s### | 1 |
|  | Healing cut on the left foot |  | s### | 1 |
|  | Temperature 37.3°C |  | s### | 1 |
| Diseases to check | Proximal and calf DVT |  |  |  |
| Diseases to exclude | Venous insufficiency |  |  |  |
|  | Underlying malignancy |  |  |  |
| Recommendations | Normal duplex ultrasound scan |  |  |  |
|  | D-dimer assay |  |  |  |

Case Citation: (18)

### Clinical Decision Support System and Explainable AI

Poor interpretability and explainability of CDSS are significant problems in medical ethics. This problem has severe and far-reaching consequences for personal and public health. (14)

The XAI for CDSS has a variety of requirements. (15), (See: S Table 3)

**Table 3.** Environments for development and execution.

|  |  |  |
| --- | --- | --- |
| Programming language | Python 3.12.x |  |
| Machine learning libraries |  |  |
| Main model | TensorFlow 2.10.x | Neural Network (NN) |
|  | TensorFlow Ranking 0.5.x | Learning To Rank (LTR) |
| Surrogate model | scikit-learn 1.2.x | k-Nearest Neighbors (k-NN) |
| Web application framework | Flask 2.3.x |  |
| JavaScript library | Vue.js 3.x |  |
| Cloud computing services | Google Cloud Platform |  |
|  | App Engine |  |
|  | Vertex AI |  |
|  | Firebase Authentication |  |

XAI research in medicine is focused on image diagnosis and less on differential diagnosis. (16)

Research on XAI with CDSS for a single disease was reported about “Type 1 diabetes + SHAPE.” (17) A little research on XAI with CDSS for multiple diseases was reported.

No research on XAI to support Evidence-Based Medicine (EBM) was reported.

This manuscript proposes the Evidence-based XAI model for CDSSs.

This model supports the physicians’ DDx with evidence.

## Design

### Objective

The aims of this article’s XAI model are as follows:

– Requests from medical professionals:

Show evidence of predicted diseases by CDSS.
– Response from the XAI:

Evidence is the case data of this medical literature.

### Technical background for XAI

The AI techniques used for implementation are as follows:

1. Example-based explanations
2. Surrogate model
3. K-Nearest Neighbors

Based on the XAI requirements for CDSS, we selected these techniques.

### Example-based explanations

Example-based explanations explain each predicted data.

The explanation is the identified instances of the training data contributing to the predicted data. (12) The features of the Example-based explanations are as follows: (12)

1. The simplest XAI method, with interpretability and explainability
2. Explain predicted data by identifying instances of the training data contributing to it
3. A mostly model-agnostic
4. Available only if a human can understand the instance’s contents The case data of CDSS is obtained from medical literature.

If it can identify instances of the training data (= case data), it can show evidence of the predicted data (= predicted diseases).

However, the Neural Network (NN) using the CDSS’s prediction model is not adapted to Example-based explanations.

### Surrogate model

The (local) surrogate model explains each predicted data of the main model. (12) The differences between the main and surrogate models are as follows: (12)

1. The main model is often uninterpretable (black box).
2. The surrogate model is an interpretable and explicable (white box) model.
3. The prediction performance of the surrogate model is lower than that of the main model.
4. The surrogate model’s predicted data is not equal to the main model.

### K-Nearest Neighbors

The features of the k-Nearest Neighbors (k-NN) are as follows:

1. The simplest AI method, with interpretability and explainability
2. Support Example-based Explanations by instances of the training data
3. Wide adaptability

a. Non-parametric supervised learning method
b. Pattern matching
c. Various distance and evaluation functions
d. High-dimensional data (ex: varieties of symptoms and diseases)
e. Scarce training data (ex: rare diseases and cases)
4. Prediction performance is lower than that of the NN

### k-Nearest Neighbors Surrogate model for Clinical Decision Support System

We propose the k-Nearest Neighbors Surrogate model (k-NN Surrogate model) as the XAI model for CDSS.

Requirements of CDSS and features of k-NN are remarkably adaptable, except for the k-NN’s prediction performance.

Requirements of XAI for CDSS and features of Example-based explanations are remarkably adaptable. The k-NN Surrogate model is a symphony of Example-based explanations, Local surrogate model, and k-NN.

It improves prediction and XAI performance by “Selecting its data closest to the main model.”

In a typical k-NN use case, the number of neighbors (*k*) is selected by hyperparameter optimization during the design phase. The value of k is fixed during the prediction phase.

In the k-NN Surrogate model, the value of k is changed for each prediction. It predicts multiple data with multiple numbers of neighbors (ex: *k* = 1 − 10). The value of closest k (*k*_*closest*) is selected by the evaluation function’s value between the main model’s predicted data and the surrogate model’s multiple predicted data. This process can closest the surrogate model’s predicted data to the main model. (defined as “Selecting” from now on)

By “Selecting,” the surrogate model’s data (the predicted data and instances of them) are good surrogates for the main model. k-NN Surrogate model can show evidence of predicted diseases to medical professionals.

### Implementation

### Medical literature, metadata, and case data

#### Medical literature

Medical literature is a source of metadata and case data. It includes medical textbooks, original articles, case reports, etc. It will be selected carefully for the CDSS’s purpose and targets. It is limited to clear sources and peer-reviewed content.

#### Metadata

Metadata is obtained from medical literature by text-mining. It includes bibliographic, disease-related, and other information.

#### Case data

Case data is obtained from the metadata by coding. It is used as training and test data for models. It includes symptoms and diseases.

Symptoms include signs and symptoms, laboratory and imaging test results, etc. Diseases include confirmed diseases, differential diseases, and their scores.

### Case ID can identify and cross-reference between the metadata and the case data. (See: Table 2, Fig 2)

### Environments for development and execution

The evaluation function for the main (NN) and surrogate (k-NN) models is the Normalized Discounted Cumulative Gain (*ndcg*).

**Fig 2.**
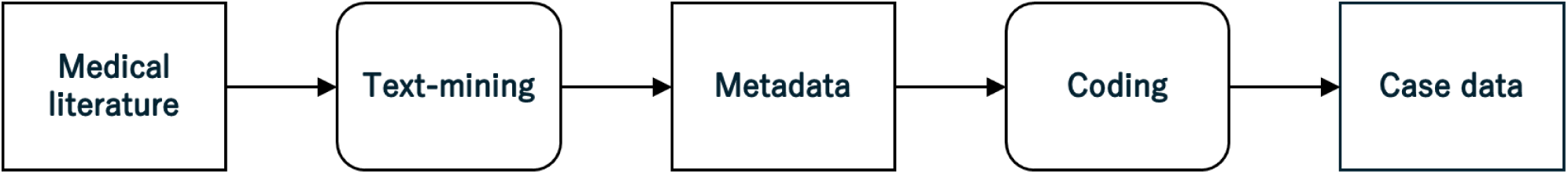
Activity diagram of text-mining and coding

The loss function for the main model (NN) is the approximate NDCG loss (*ANDCG*). (19) (See: Table 3, Table 4)

**Table 4.** Configurations of models.

| Models | Methods | Items | Values, etc. |
| --- | --- | --- | --- |
| Main | Neural Network (NN) | Input layer | 583 |
| | | Hidden layer | $1 \times 1024$ |
|  |  | Output layer | 1000 |
|  |  | Loss functions | *ANDCG* |
|  |  | Evaluation functions | *ndcg, ndcg@k* |
| Surrogate | k-Nearest Neighbors (k-NN) | Number of neighbors | 1 – 10 |
|  |  | Distance function | *Kendall's W* |
| | | Weight function | $1/\text{distance}$ |
|  |  | Evaluation functions | *ndcg, ndcg@k* |

### Training of the models

The main and surrogate models use same datasets. (See: Fig 3)

**Fig 3.**
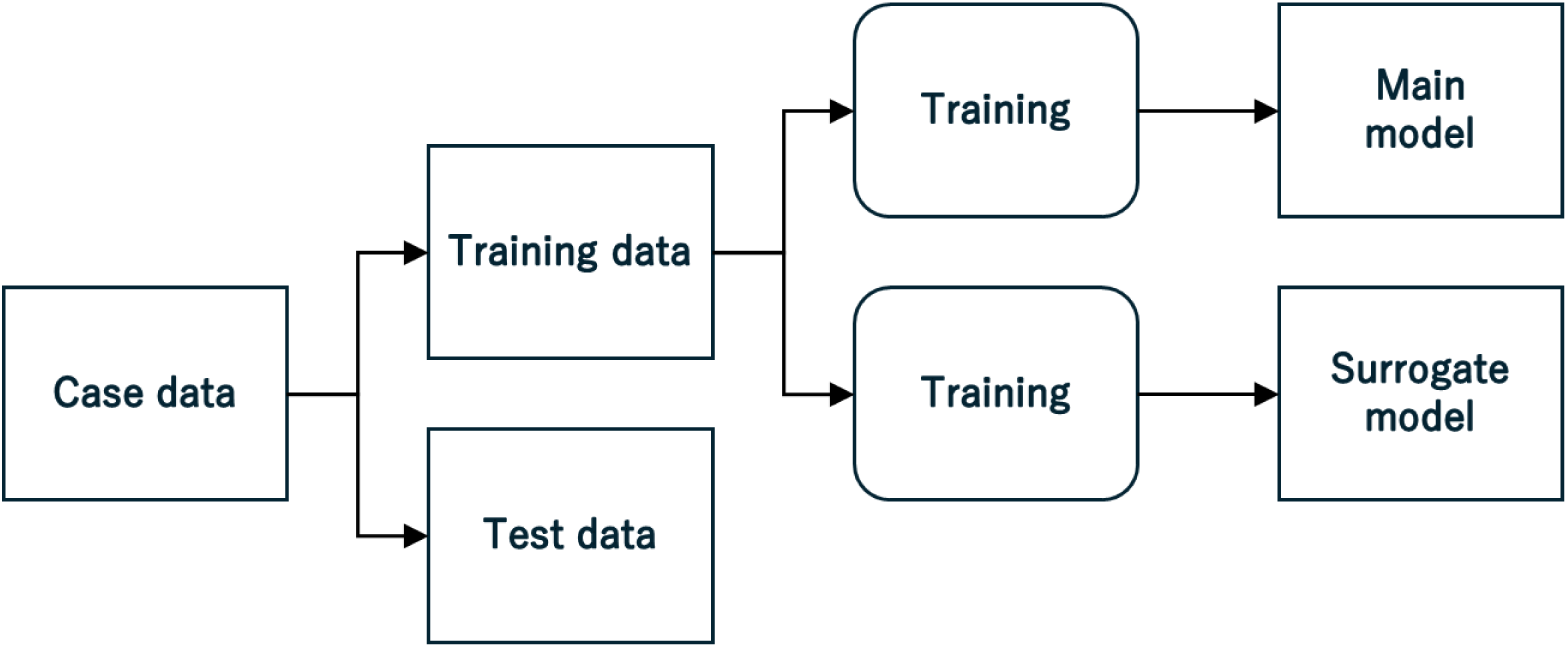
Activity diagram of training (See: S Code 1)

### Prediction and XAI models (See: Fig 4, Fig 5)

### Evaluation

Datasets for evaluation use k-fold cross-validation of case data. (See: Fig 6)

**Fig 4.**
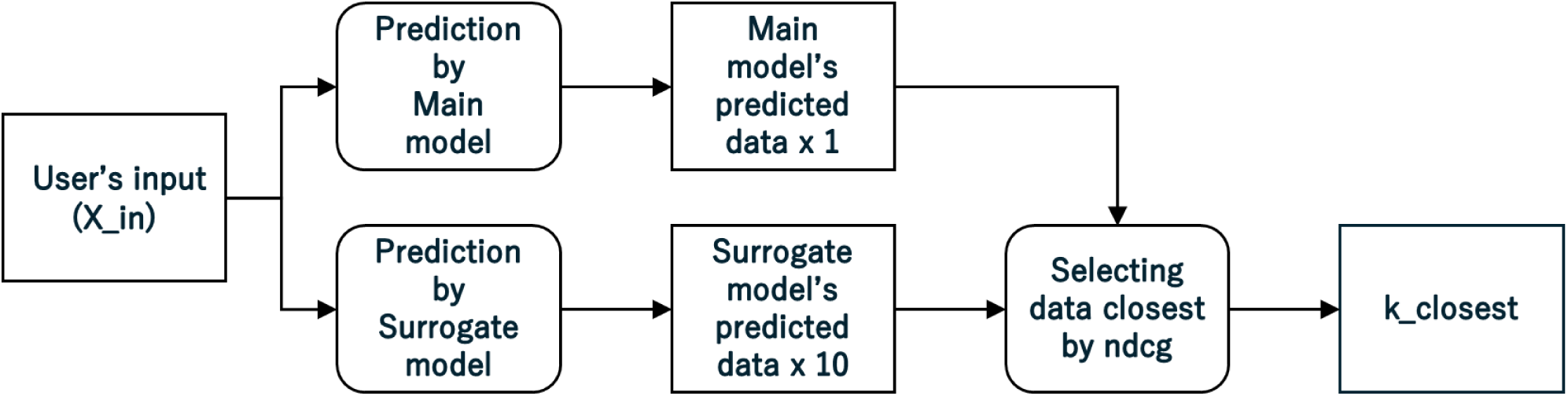
Activity diagram of prediction (See: S Code 1)

**Fig 5.**
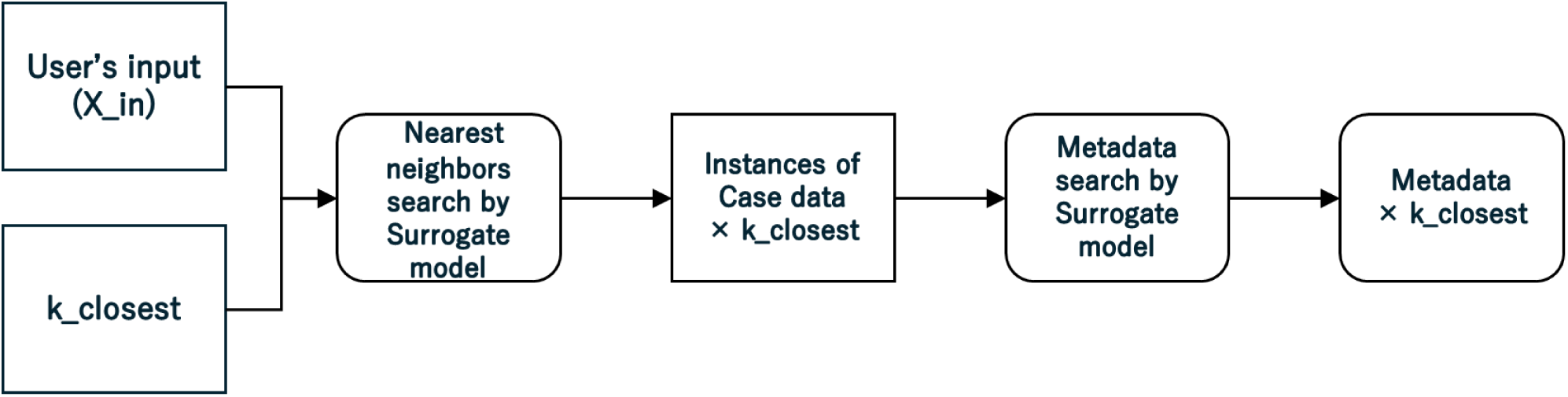
Activity diagram of explanation (See: S Code 1)

**Fig 6.**
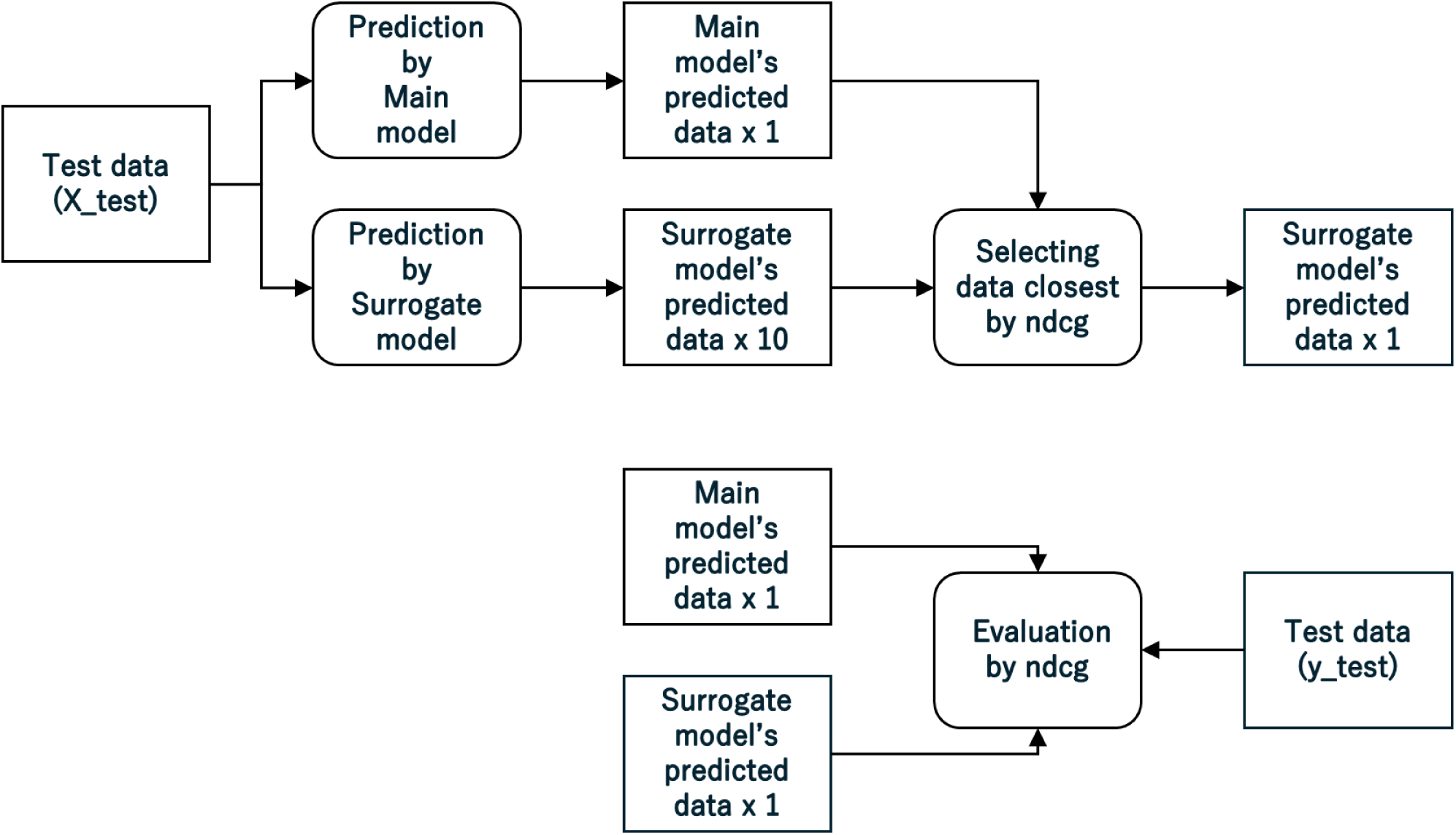
Activity diagram of evaluation (See: S Code 1)

## Results and Discussion

### Prediction performance

The prediction performance of the main and surrogate models was evaluated. (See: Table 5)

**Table 5.** Prediction performance of the main and surrogate models)

| Models | Data | *k* | *ndcg* | *ndcg@1* | *ndcg@3* | *ndcg@5* | *ndcg@10* |
| --- | --- | --- | --- | --- | --- | --- | --- |
| Main | *ndcg_nn* | ---- | 0.6684 | 0.4609 | 0.5393 | 0.5690 | 0.5988 |
| Surrogate | *ndcg_k1[]* | 1 | 0.5348 | 0.3417 | 0.3782 | 0.3986 | 0.4231 |
|  |  | 2 | 0.5636 | 0.3444 | 0.4122 | 0.4366 | 0.4632 |
|  |  | 3 | 0.5738 | 0.3554 | 0.4201 | 0.4467 | 0.4754 |
|  |  | 4 | 0.5784 | **0.3597** | **0.4242** | **0.4517** | 0.4810 |
|  |  | 5 | 0.5791 | 0.3587 | 0.4240 | 0.4514 | **0.4812** |
|  |  | 6 | **0.5791** | 0.3581 | 0.4222 | 0.4510 | 0.4812 |
|  |  | 7 | 0.5778 | 0.3526 | 0.4206 | 0.4484 | 0.4797 |
|  |  | 8 | 0.5772 | 0.3510 | 0.4190 | 0.4480 | 0.4788 |
|  |  | 9 | 0.5756 | 0.3487 | 0.4165 | 0.4451 | 0.4766 |
|  |  | 10 | 0.5735 | 0.3440 | 0.4137 | 0.4421 | 0.4740 |
|  | *ndcg_k1_closest* | ---- | 0.6264 | 0.4361 | 0.4930 | 0.5158 | 0.5385 |

The “standalone” surrogate model’s prediction performance is lower than the main model.
When using “Selecting,” the surrogate model's prediction performance is higher than the “standalone” surrogate model.
However, it is lower than the main model.

### XAI performance

The XAI performance of the surrogate model was evaluated. (See: Table 6)

**Table 6.** XAI performance of the surrogate model.

| Model | Data | *k* | *ndcg* | *ndcg*@1 | *ndcg*@3 | *ndcg*@5 | *ndcg*@10 |
| --- | --- | --- | --- | --- | --- | --- | --- |
| Surrogate | *ndcg_k2[]* | 1 | 0.8049 | 0.6526 | 0.5975 | 0.5900 | 0.5994 |
|  |  | 2 | 0.8255 | 0.6775 | 0.6521 | 0.6415 | 0.6429 |
|  |  | 3 | 0.8335 | 0.6912 | 0.6674 | 0.6601 | 0.6600 |
|  |  | 4 | 0.8381 | 0.7016 | 0.6763 | 0.6690 | 0.6698 |
|  |  | 5 | 0.8403 | **0.7040** | 0.6800 | 0.6731 | 0.6748 |
|  |  | 6 | 0.8414 | 0.7033 | 0.6812 | 0.6755 | 0.6775 |
|  |  | 7 | 0.8421 | 0.7038 | 0.6828 | 0.6770 | 0.6790 |
|  |  | 8 | 0.8424 | 0.7019 | **0.6831** | 0.6775 | 0.6796 |
|  |  | 9 | **0.8426** | 0.7012 | 0.6829 | **0.6776** | **0.6801** |
|  |  | 10 | 0.8422 | 0.6979 | 0.6812 | 0.6768 | 0.6798 |
|  | *ndcg_k2_closest* | ---- | 0.8800 | 0.8654 | 0.8101 | 0.7810 | 0.7566 |

XAI performance is the similarity between the predicted data of the main and surrogate models. The similarity is evaluated by the evaluation function.
The “standalone” surrogate model’s XAI performance is high.
When using “Selecting,” the surrogate model’s XAI performance is higher than the “standalone” surrogate model.
“Selecting data closest to the main model” improves the surrogate model’s prediction and XAI performance.

## Conclusions

The k-NN Surrogate model is a useful XAI model for CDSS.

It is a symphony of Example-based explanations, Local surrogate model, and k-NN.

k-NN is adapted to high-dimensional data (ex: varieties of symptoms and diseases) and scarce case data (ex: rare diseases and cases).

Our CDSS’s case data are the same as the evidence in medical literature.

For CDSSs with similar aims and features, Example-based explanations are helpful and easy to implement. The k-NN Surrogate model is an Evidence-based XAI for CDSS Uncertainty Quantification (UQ) is an essential issue for AI and CDSS. (20), (21) k-NN is adapted to Conformal Prediction (CP), one of UQ’s. (22), (23) The k-NN Surrogate model will also contribute to these improvements in performance.

Unlike current commercial Large Language Models (LLMs), Our CDSS shows evidence of the predicted diseases to medical professionals.

This emphasis on evidence provides a sense of reassurance and confidence in the system’s capabilities. It is important to remember that XAI and the k-NN Surrogate model are beneficial and can change the game for CDSSs.

## Additional Information

## Supporting information

S Table 1 Example of predicted diseases

S Table 2 List of Explainable Artificial Intelligence methods

S Table 3 Requirements of Explainable Artificial Intelligence for Clinical Decision Support System

S Code 1 Pseudo code for k-Nearest Neighbors Surrogate model

## Acknowledgements

Not applicable.

## Author Approval

This manuscript has been seen and approved by all listed authors. This manuscript has not been accepted or published by a journal.

## Competing Interests

The authors have declared no competing interest.

## Declarations

1. I have the right to post this manuscript and confirm that all authors have assented to posting of the manuscript and inclusion as authors.
2. I confirm all relevant ethical guidelines have been followed, and any necessary IRB and/or ethics committee approvals have been obtained. The study does not describe the use of any human data, samples, or any research involving human subjects.
3. I confirm that all necessary patient/participant consent (including consent to publish) has been obtained and the appropriate institutional forms have been archived. I confirm that any patient/participant/sample identifiers included were not known to anyone (e.g., hospital staff, patients or participants themselves) outside the research group so cannot be used to identify individuals.
4. I understand that all clinical trials and any other prospective interventional studies must be registered with an ICMJE-approved registry, such as ClinicalTrials.gov. I confirm that any such study reported in the manuscript has been registered and the trial registration ID is provided.
5. I am legally responsible for the content of the article.
6. I have followed all appropriate research reporting guidelines, such as any relevant EQUATOR Network research reporting checklist(s) and other pertinent material, if applicable.

## Data Availability Statement and Links

All data produced in the present study are available upon reasonable request to the authors. https://www.diagnosis.or.jp/

## Funding Statement

This study did not receive any funding.

## Supporting information

1. S Table 1 Example of predicted diseases

a. Filename: S_table_01_Example_of_the_predicted_diseases.pdf
b. Case citation: (6)
c. Notes: The predicted diseases of the Fig 1.
2. S Table 2 List of Explainable Artificial Intelligence methods

a. Filename: S_table_02_List_of_XAI_methods.pdf
b. Citation: (12), (13)
3. S Table 3 Requirements of Explainable Artificial Intelligence for Clinical Decision Support System

a. Filename: S_table_03_Requirements_of_XAI_for_CDSS.pdf
b. Citation: (15)
4. S Code 1 Pseudo code for k-Nearest Neighbors Surrogate model

a. Filename: S_code_01_Pseudo_code.pdf
b. Notes1: The code of the activity diagrams (Fig 3, Fig 4, Fig 5, Fig 6).
c. Notes2: No guarantee of executability.

End

## Notes

### Summary of Updates

Fixing errors in figure and table references. Polishing the text.

## References

1. Sutton RT, Pincock D, Baumgart DC, Sadowski DC, Fedorak RN, Kroeker KI. An overview of clinical decision support systems: benefits, risks, and strategies for success. NPJ Digit Med [Internet]. 2020 [cited 2024 Jul 18];3(1). Available from: https://www.nature.com/articles/s41746-020-0221-y

2. Miyachi Y, Ishii O, Torigoe K. Design, implementation, and evaluation of the computer-aided clinical decision support system based on learning-to-rank: collaboration between physicians and machine learning in the differential diagnosis process. BMC Med Inform Decis Mak [Internet]. 2023 [cited 2024 Jul 18];23(1). Available from: https://bmcmedinformdecismak.biomedcentral.com/articles/10.1186/s12911-023-02123-5

3. Miyachi Y, Ishii O, Torigoe K. Can AI Clinical Decision Support System show evidence?: Explainable AI in collaboration between Neural Network and Surrogate model for Learning to Rank. In: The 43rd Joint Conference on Medical Informatics [Internet]. 2023 [cited 2024 Jul 6]. Available from: https://jglobal.jst.go.jp/detail?JGLOBAL_ID=202402210603144321

4. Miyachi Y, Ishii O, Torigoe K. Can AI Clinical Decision Support System show Evidence and Humility?: Fusion of XAI and UQ with Surrogate model. In: The 38th Annual Conference of the Japanese Society for Artificial Intelligence [Internet]. 2024 [cited 2024 Jul 18]. Available from: https://www.jstage.jst.go.jp/article/pjsai/JSAI2024/0/JSAI2024_2A1GS1004/_article/-char/en

5. The Society for Computer-aided Clinical Decision Support System. DiagnosticNN on the Web [Internet]. [cited 2023 Aug 6]. Available from: https://www.diagnosis.or.jp/

6. Kratka A, Tedrow UB, Mitchell RN, Miller AL, Loscalzo J. A Stormy Heart. New England Journal of Medicine [Internet]. 2023 [cited 2024 Jul 18];388(1). Available from: https://www.nejm.org/doi/full/10.1056/NEJMcps2116690

7. Isabel Healthcare. Isabel Healthcare: Differential Diagnosis Tool [Internet]. [cited 2024 Jul 6]. Available from: https://www.isabelhealthcare.com/

8. BraineHealth. Diagnosio | Intelligent healthcare for all [Internet]. [cited 2024 Jul 6]. Available from: https://www.diagnosio.com/

9. The Japanese Society of Internal Medicine. J-CaseMap [Internet]. [cited 2023 Aug 6]. Available from: https://www.naika.or.jp/j-casemap/

10. Database Center for Life Science. PubCaseFinder [Internet]. [cited 2023 Aug 6]. Available from: https://pubcasefinder.dbcls.jp/

11. Gunning D, Aha DW. DARPA’s explainable artificial intelligence program. AI Mag [Internet]. 2019 [cited 2024 Jul 18];40(2). Available from: https://onlinelibrary.wiley.com/doi/full/10.1609/aimag.v40i2.2850

12. Molnar C. Interpretable Machine Learning: A Guide for Making Black Box Models Explainable [Internet]. [cited 2023 Aug 6]. Available from: https://christophm.github.io/interpretable-ml-book/

13. Burkart N, Huber MF. A survey on the explainability of supervised machine learning [Internet]. Vol. 70, Journal of Artificial Intelligence Research. 2021 [cited 2024 Jul 18]. Available from: https://dl.acm.org/doi/10.1613/jair.1.12228

14. Amann J, Blasimme A, Vayena E, Frey D, Madai VI. Explainability for artificial intelligence in healthcare: a multidisciplinary perspective. BMC Med Inform Decis Mak [Internet]. 2020 [cited 2024 Jul 18];20(1). Available from: https://bmcmedinformdecismak.biomedcentral.com/articles/10.1186/s12911-020-01332-6

15. Ploug T, Holm S. The four dimensions of contestable AI diagnostics-A patient-centric approach to explainable AI. Artif Intell Med [Internet]. 2020 [cited 2024 Jul 18];107. Available from: https://www.sciencedirect.com/science/article/pii/S0933365720301330

16. National Library of Medicine. PubMed [Internet]. [cited 2024 Jul 6]. Available from: https://pubmed.ncbi.nlm.nih.gov/

17. Prendin F, Pavan J, Cappon G, Del Favero S, Sparacino G, Facchinetti A. The importance of interpreting machine learning models for blood glucose prediction in diabetes: an analysis using SHAP. Sci Rep [Internet]. 2023 [cited 2024 Jul 18];13(1). Available from: https://www.nature.com/articles/s41598-023-44155-x

18. Stern S, Cifu A, Altkorn D. Symptom to Diagnosis [Internet]. Vol. 1, McGraw-Hill Education. 2015 [cited 2024 Jul 10]. Available from: https://www.mheducation.com/highered/product/symptom-diagnosis-evidence-based-guide-fourth-edition-stern-altkorn/9781260121117.html

19. Bruch S, Zoghi M, Bendersky M, Najork M. Revisiting approximate metric optimization in the age of deep neural networks. In: SIGIR 2019 - Proceedings of the 42nd International ACM SIGIR Conference on Research and Development in Information Retrieval [Internet]. 2019 [cited 2024 Jul 18]. Available from: https://dl.acm.org/doi/10.1145/3331184.3331347

20. Seoni S, Jahmunah V, Salvi M, Barua PD, Molinari F, Acharya UR. Application of uncertainty quantification to artificial intelligence in healthcare: A review of last decade (2013–2023) [Internet]. Vol. 165, Computers in Biology and Medicine. 2023 [cited 2024 Jul 18]. Available from: https://www.sciencedirect.com/science/article/pii/S001048252300906X

21. Vazquez J, Facelli JC. Conformal Prediction in Clinical Medical Sciences [Internet]. Vol. 6, Journal of Healthcare Informatics Research. 2022 [cited 2024 Jul 18]. Available from: https://link.springer.com/article/10.1007/s41666-021-00113-8

22. Papadopoulos H, Vovk V, Gammerman A. Regression conformal prediction with nearest neighbours. Journal of Artificial Intelligence Research [Internet]. 2011 [cited 2024 Jul 18];40. Available from: 10.1613/jair.3198

23. Boström H, Johansson U, An Nguyen K, Luo Z, Carlsson L. crepes: a Python Package for Generating Conformal Regressors and Predictive Systems [Internet]. Vol. 179, Proceedings of Machine Learning Research. 2022 [cited 2024 Jul 10]. Available from: https://proceedings.mlr.press/v179/bostrom22a.html

