## Supplementary material for "Evidence-based XAI of clinical decision support systems for differential diagnosis: Design, implementation, and evaluation": S Table 1 Example of predicted diseases

| | Inputted symptoms |
|---|---|
| a | heart failure (right and/or left); BNP/NT-proBNP increase |
| b | CRP, positive; C-reactive protein, positive |
| c | ESR elevation; erythrocyte sedimentation rate elevation |
| d | pericardial fluid retention |
| e | lymphadenopathy/adenopathy (take no account of the property/location) |
| f | ANA (anti-nuclear antibody), positive (320 times or more) |
| g | thickening/swelling in any part of the body in image views |
| h | any abnormal changes/lesions in the chest radiograph |
| i | palpitation; fast heartbeat; abnormally strong heartbeat |
| j | confusion; lightheadedness; decreased alertness |
| k | shortness of breath; gasping; irregular respiration; exertional dyspnea |
| l | chest tightness in the chest; feel oppressed in the chest |
| m | hypertension; have a history of hypertension |
| n | ECG (electrocardiogram) abnormalities (take no account of any kinds) |
| o | tachycardia (almost 100 per minute or more) |
| p | increased CPK/CK or CK-MB (CK; creatin kinase); increased troponin T |

| Ranking | predicted diseases |
| --- | --- |
| 1 ◎ | **myocarditis** (note: all types such as viral/**giant cell**/eosinophil/lymphocytic/hypersensitivity) |
| ▼ | fulminant myocarditis |
| ▼ | myocardial pericarditis |
| ▼ | drug-induced myocarditis (side effects of salazopyrine, etc.)/drug-induced myocardial damage (e.g., adriamycin) |
| ▼ | cocaine myocarditis |
| ▼ | tick-borne illnesses |
| 2 ◎ | acute epicarditis |
| ◎ | acute pericarditis |
| ◎ | acute myocarditis |
| ◎ | acute myopericarditis (various causes/be careful about enterovirus, coxsackievirus, tuberculosis, malignant tumor, etc.) |
| ◎ | uremic epicarditis |
| ▼ | Dressler syndrome/post-cardiac injury/post-traumatic pericarditis |
| ▼ | intramyocardial abscess |
| ▼ | extrapericardial fat necrosis |
| 3 ◎ | cardiac tamponade/cardiogenic shock (cardiac rupture, aortic injury, aortic dissection/thromboembolic type aortic dissection (either surgery or conservative therapy), (cancerous/tuberculous) pericarditis, consider causes such as bleeding tendency, etc.) sepsis-induced cardiomyopathy or obstructive shock (caused by tension pneumothorax, (giant) esophageal hiatal hernia, severe pulmonary infarction, tension pneumo-mediastinum, etc.) |
| ◎ | isolated persistent left superior vena cava |
| ▼ | pericardial tumors |

| Ranking | predicted diseases |
|---|---|
| 4 ◎ | cardiac amyloidosis (the most often derived from amyloid light chain (AL) associated with blood disorders) |
| ▼ | wild type ATTR (transthyretin) amyloidosis (senile systemic amyloidosis) |
| 5 ◎ | right ventricular infarction (pure right ventricular infarction of the right ventricle is about 3.3% of myocardial infarction) |
| ▼ | right heart failure |
| ▼ | double outlet right ventricle |
| ▼ | corrected transposition of the great arteries |
| ▼ | and persistent truncus arteriosus |
| ▼ | complications of left ventricular assisted artificial heart (gastrointestinal hemorrhage, right heart failure, pump thrombosis, ejection system infection, etc.) |
| 6 ◎ | sarcoidosis (including Heerfordt's syndrome) |
| ▼ | ((multiple) reticulohistiocytosis, foreign body (beryllium, silica) granuloma, IgG4-related disease should be kept in mind) |
| 7 ◎ | Catecholamine-induced cardiomyopathy (also remind/consider catecholamine secreting glomus tumor, pheochromocytoma, etc.) |
| ◎ | sometimes stress cardiomyopathy/heart attack syndrome/takotsubo cardiomyopathy |
| 8 ◎ | pheochromocytoma/pheochromocytoma crisis |
| ▼ | (malignant, familial) paraganglioma (para-aortic (70%), Zuckerkandl body, cervical, mediastinum, bladder neck, etc.) |
| ▼ | hereditary pheochromocytoma/paraganglioma syndrome (HPPS) |
| ▼ | pheochromocytoma-related syndrome |
| ▼ | primary pigmented nodular adrenocortical disease (PPNAD) |
| ▼ | (benign, malignant) carotid body tumor |
| 9 ◎ | thyroid crisis |

| Ranking | predicted diseases |
|---|---|
| 10 ◎ | pulmonary arterial hypertension (familial/idiopathic) |
| ▼ | pulmonary arterial hypertension associated with connective tissue diseases |
| ▼ | pulmonary valve stenosis (regurgitation) |
| ▼ | persistent pulmonary hypertension of the newborn (PPHN) |
| ▼ | pregnancy-related (postpartum) pulmonary arterial hypertension |

◎: Common diseases

▼: Rare diseases

Case citation:

Kratka A, Tedrow UB, Mitchell RN, Miller AL, Loscalzo J. A Stormy Heart. New England Journal of Medicine. 2024; 388(1).
Available from: https://www.nejm.org/doi/full/10.1056/NEJMcps2116690
