## Supplementary material for "Evidence-based XAI of clinical decision support systems for differential diagnosis: Design, implementation, and evaluation": S Table 2 List of Explainable Artificial Intelligence methods

| Types | Methods |
| --- | --- |
| Interpretable Machine Learning | Linear Regressions |
| | Logistic Regressions |
| | Generalized Linear Models (GLMs) |
| | Generalized Additive Models (GAMs) |
| | Decision Tree |
| | Decision Rules |
| | RuleFit |
| | Naive Bayes Classifier |
| | **K-Nearest Neighbors (k-NN)** |
| Example-Based Explanations | Counterfactual explanations |
| | Adversarial examples |
| | Prototypes and Criticisms |
| | Influential Instances |
| | Deletion Diagnostics |
| | Influence Functions |
| | **K-Nearest Neighbors (k-NN)** |

| Types | Methods |
| --- | --- |
| Global Model-Agnostic Methods | Interpretable Machine Learning |
| | Partial Dependence Plot (PDP) |
| | Accumulated Local Effects (ALE) Plot |
| | Feature Interaction |
| | Functional Decomposition |
| | Permutation Feature Importance (PFI) |
| | Global Surrogate models |
| | Prototypes and Criticisms |
| Local Model-Agnostic Methods | Interpretable Machine Learning |
| | Example-Based Explanations |
| | Partial Dependence (PD) |
| | Individual Conditional Expectation (ICE) |
| | Local Surrogate models |
| | Decision tree surrogate model |
| | **k-NN Surrogate Model** [*1)] |
| | Local Interpretable Model-agnostic Explanations (LIME) |
| | Counterfactual Explanations |
| | Scoped Rules (Anchors) |
| | Shapley Values |
| | SHapley Additive exPlanations (SHAP) |

| Types | Methods |
| --- | --- |
| Neural Network Interpretation | Learned Features |
| | Pixel Attribution (Saliency Maps) |
| | Detecting Concepts |
| | Adversarial examples |
| | Influential Instances |
| | Deletion Diagnostics |
| | Influence Functions |

*1: Add by authors

Citation:

Molnar C. Interpretable Machine Learning: A Guide for Making Black Box Models Explainable.
Available from: https://christophm.github.io/interpretable-ml-book/

Burkart N, Huber MF. A survey on the explainability of supervised machine learning. Journal of Artificial Intelligence Research.
2021; Vol. 70. Available from: https://dl.acm.org/doi/10.1613/jair.1.12228
