## Supplementary material for "Evidence-based XAI of clinical decision support systems for differential diagnosis: Design, implementation, and evaluation": S Table 3 Requirements of Explainable Artificial Intelligence for Clinical Decision Support System

| Types | | Questions | Required Explanations |
|---|---|---|---|
| Data | Types of data | What types of personal data are used? | The decision D was made on the basis of data of type X, Y, Z about you. |
| | Data sources | Where do your data come from? | The decision D was based on data from sources X, Y, Z. |
| Bias | Training data | On which data was the AI trained? | The decision D was made by a system trained on existing data of type X, Y, Z. |
| | Tagging groups | Who tagged training data? | The decision D was made by a system trained on data tagged by X, Y, Z. |
| | Tested for bias | Was training data or AI system tested for bias? | The decision D was made by a system tested for bias of type X, Y, Z. |
| Diagnostic Performance | Performance | What is the performance of the AI system? | The decision D was made by a system with a performance of X, Y, Z. |
| | Performance testing | How was the performance determined? | The decision D was made by a system with a performance determined by tests X,Y, Z. |
| | Essential indicators | What are key variables of AI decision-making? | The key input data resulting in decision D was X, Y, Z. |
| | Alternatives | Are alternatives considered? | The alternatives to decision D are X, Y, Z with a probability of x, y, z (< d). |
| | Longevity | When is decision reconsidered? | The decision D will be reconsidered if conditions X, Y, Z obtain. |
| Decision | AI involvement | To what degree is AI making the decision? | The decision D involved an AI system with respect to X, Y, Z. |
| | Human involvement | To what degree are humans making the decision? | The decision D was wholly/partly made by health professionals X, Y, Z. |
| | Responsibility | Who is responsible for the decision? | The objective/legal responsibility for decision D is held by X, Y, Z. |
| | **Evidence** [*2)] | **What is the evidence on which decision D was based?** | **The decision D was based on instances X, Y, Z of the training data.** |

*2: Add by authors

Citation:

Ploug T, Holm S. The four dimensions of contestable AI diagnostics- A patient-centric approach to explainable AI. Artif Intell Med. 2020;107.

Available from: https://www.sciencedirect.com/science/article/pii/S0933365720301330
