## Supplementary material for "Evidence-based XAI of clinical decision support systems for differential diagnosis: Design, implementation, and evaluation": S Code 1 Pseudo code for k-Nearest Neighbors Surrogate model

```
# Pseudo code for k-Nearest Neighbors Surrogate model

# No guarantee of executability.

################################################################################

# Training of the main and surrogate models with the case data

# Main model

main_model.fit(X_train, y_train)

# Surrogate model

surrogate_model.fit(X_train, y_train)

################################################################################

# Prediction and explanation by the main and surrogate models with user's input

max_n_neighbors = 10

# Main model

y_pred = main_model.predict(X_in)

# Surrogate model

ndcg_k2_list = list()

for j in range(max_n_neighbors):
    k = j + 1
    y_pred_k = surrogate_model.predict(X_in, k)
    ndcg_k2 = ndcg(y_pred, y_pred_k)
    ndcg_k2_list.append(ndcg_k2)

k_closest = argmax(ndcg_k2_list) + 1
ndcg_k2_closest = ndcg_k2_list[k_closest - 1]

(neigh_dist, neigh_ind) = surrogate_model.kneighbors(X_in, k_closest)

meta_data_list = surrogate_model.search_meda_data(neigh_ind)

################################################################################

# Evaluation of the main and surrogate models with the case data

n_tests = 6000
max_n_neighbors = 10

for i in range(n_tests):
    X_in = X_test[i]
    y_true = y_test[i]

    # Main model

    ndcg_nn = ndcg(y_true, y_pred)

    # Surrogate model

    ndcg_k1_list = list()
    ndcg_k2_list = list()
    for j in range(0, max_n_neighbors):
        k = i + 1
        y_pred_k = surrogate_model.predict(X_in, k)
        ndcg_k1 = ndcg(y_true, y_pred_k)
        ndcg_k2 = ndcg(y_pred, y_pred_k)
        ndcg_k1_list.append(ndcg_k1)
        ndcg_k2_list.append(ndcg_k2)

    k_closest = argmax(ndcg_k2_list) + 1
    ndcg_k1_closest = ndcg_k1_list[k_closest - 1]
    ndcg_k2_closest = ndcg_k2_list[k_closest - 1]
```
